# Covid-19 Vaccine Acceptance Among People Incarcerated in Connecticut State Jails

**DOI:** 10.1101/2022.05.19.22275339

**Authors:** Margaret L. Lind, Byron S. Kennedy, Murilo Dorion Nieto, Amy J. Houde, Peri Sosensky, Ryan Borg, Derek A.T. Cummings, Albert I. Ko, Robert P. Richeson

## Abstract

**Objective:** To assess the Connecticut Department of Correction’s (DOC) COVID-19 vaccine program within jails.

**Methods:** We conducted a retrospective cohort analysis among people who were incarcerated in a DOC-operated jail between February 2 and November 8, 2021, and were eligible for vaccination at the time of incarceration (intake). We compared the vaccination rates before and after incarceration using an age-adjusted survival analysis with a time-varying exposure of incarceration and an outcome of vaccination.

**Results:** During the study period, 3,716 people spent ≥1 night in jail and were eligible for vaccination at intake. Of these residents, 136 were vaccinated prior to incarceration, 2,265 had a recorded vaccine offer, and 476 were vaccinated while incarcerated. The age-adjusted hazard of vaccination following incarceration was significantly higher than prior to incarceration (12.5; 95% CI: 10.2-15.3).

**Conclusions:** We found that residents were more likely to become vaccinated in jail than the community. Though these findings highlight the utility of vaccination programs within jails, the low level of vaccination in this population speaks to the need for additional program development within jails and the community.

## Introduction

COVID-19 has disproportionately affected people who experience incarceration in state or federal run correctional facilities.^1–5^ To mitigate transmission, US state and federal Department of Corrections (DOCs) implemented vaccine programs in the winter of 2020-2021.^2,6,7^ However, vaccine hesitancy is common among incarcerated populations due to distrust of the medical community and uncertainty around vaccine effectiveness.^7^ Consequently, data from state and federal correctional facilities suggest vaccine acceptance within residents ranges between 40-67%.^6–10^

Despite the continued need for vaccine programs within correctional facilities, the impact vaccine programs have on overall vaccine acceptance within people who experience incarceration is not well characterized, especially among jailed populations. Herein we leveraged data from Connecticut-DOC-operated jails to evaluate the success of the DOC’s vaccination program within jails. Specifically, we calculated vaccine acceptance among residents with recorded vaccine offers. Further, to test if the program resulted in increased vaccination within this population, we compared vaccination rates among newly incarcerated people before and after incarceration.

## Methods

The Connecticut DOC initiated a COVID-19 vaccination program on February 2, 2021 for residents of their 12 prisons (long-term post sentencing facilities) and five jails (pre or short-term post sentencing facilities).^8^ Under the program, vaccination was offered to residents who qualified for vaccination according to state-defined eligibility (e.Table1). Residents who were partially vaccinated prior to incarceration were offered second doses. Anonymized movement, demographic, community vaccination history, and within-facility vaccine offer and acceptance data were extracted from DOC databases. From these data, we identified people who spent at least one night in a DOC-operated jail while vaccine eligible between February 2 and November 8, 2021. We limited our analysis to residents whose first incarceration during the study period was in jail and excluded data collected after a resident’s first discharge from jail. From these data, we estimated vaccine acceptance as the number of accepted doses over the number of recorded, offered doses.

To compare vaccination rates before and after incarceration, we restricted our sample to residents who were vaccine eligible at the time of incarceration (intake), had not been vaccinated prior to February 2, 2021, and were incarcerated on or after February 2, 2021. We performed a survival analysis with a time varying exposure of incarceration and an outcome of vaccination (receipt of a first dose). Follow-up time was defined as the interval between vaccine eligibility within the DOC (e.Table1) and the administration of the first vaccine dose, death, or first discharge from jail, whichever occurred first. We visualized the difference in the probability of vaccination using Kaplan Meier curves and estimated age-adjusted hazard ratios (HR) using Cox Proportional Hazards models. Additionally, we generated race/ethnicity-specific (non-Hispanic Black, non-Hispanic White, Hispanic) Kaplan Meier curves and age-adjusted HRs. This work was determined to be a public health surveillance activity by the Yale University Institutional Review Board.

We conducted four sensitivity analyses testing the robustness of our findings to alternative inclusion criteria. Specifically, we conducted analyses that included community vaccinations prior to February 2, restricted to residents vaccinated in the community or with recorded vaccine offers, restricted to residents who were vaccine eligible in the community for at least one week, and restricted to residents incarcerated on or after May 15 (Supplement).

## Results

Between February 2 and November 8, 2021, 6,522 people stayed ≥1 night in a DOC-operated jail while vaccine eligible and had not been previously incarcerated during the study period (median length of stay: 70 days; Interquartile Range [IQR]: 13-182 days). Among the 6,358 residents who were unvaccinated at intake, 4,980 (78.3%) had a recorded vaccine offer (e.Figure1). Of the 4,817 residents whose first recorded offer occurred in jail, 24.9% (1,198/4,817) accepted after the first and 18.3% (664/3,619) accepted following subsequent recorded offers, resulting in an overall acceptance proportion of 38.7% (1,862/4,817). Vaccine acceptance was higher for non-Hispanic Whites and Hispanics than non-Hispanic Blacks and for residents incarcerated at the time of program rollout (February 2, 2021; e.Table2).

Of the 3,716 residents who were eligible for vaccination at intake (e.Figure2), 136 (3.7%) were vaccinated prior to incarceration, 2,265 (61.0%) had a recorded offer and 476 (12.8%) became vaccinated while in jail. Residents spent more time eligible for vaccination in the community (79 days [IQR: 41-183]) than in jail (14 days [IQR: 3-31]; e.Table3).

The probability of vaccination was higher following incarceration than prior to incarceration across all residents and among non-Hispanic Black, non-Hispanic White, and Hispanic residents (Figure1A). Following adjustment for age, the hazard of vaccination was 12.5 (95% Confidence Intervals [CI]: 10.2-15.3) times higher following incarceration than prior to incarceration. The HR was highest for non-Hispanic Whites (14.2 [CI: 10.1-20.0]) and lowest for non-Hispanic Blacks (10.6 [CI: 7.5-14.9]; Figure1B). Global Schoenfeld Residual pvalues were >0.05 (e.Figures3-6). ^6^

**Figure 1:**
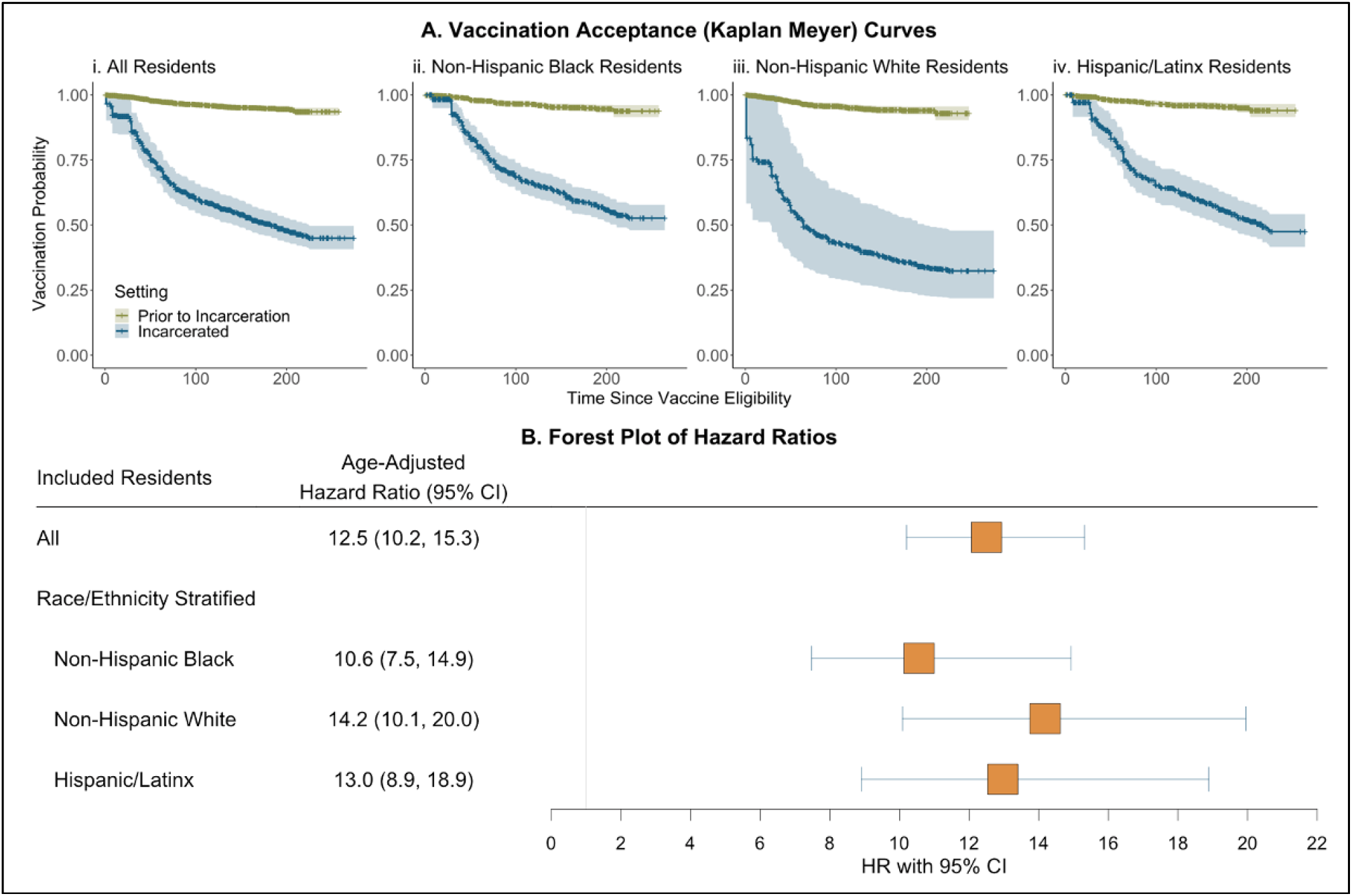
Vaccination Within Newly Incarcerated Residents of Connecticut State Operated Jails Before and After Incarceration. (A) Kaplan Meier curves displaying the probability of vaccination among newly incarcerated residents of Connecticut State jails who were eligible for vaccination on the day of their incarceration. Plots are faceted by included resident population: (i) All residents, (ii) Non-Hispanic Black residents, (iii) Non-Hispanic White residents, (iv) Hispanic/Latinx residents. (B) Forest plot of age-adjusted hazard ratios compare instantaneous vaccination before and after incarceration. Vaccine eligibility was based on the individual’s birthdate and the date that their respective age group was eligible to receive the vaccine.

The age-adjusted HRs were significant for each sensitivity analysis (e.Tables4-7). The HR was highest when we restricted to residents incarcerated on or after May 15 (22.2, e.Table4) and lowest when we restricted to residents vaccinated in the community or with recorded vaccine offers (7.0, e.Table5). For each examined scenario, the race stratified HR for non-Hispanic Blacks had the lowest HR (range: 5.5-18.6; e.Tables4-7).

## Discussion

Vaccine programs were implemented within correctional facilities to help mitigate the risk of COVID-19 among residents.^2,6,7^ In our evaluation of the Connecticut DOC’s vaccine program within jails, we observed a slightly lower vaccine acceptance proportion than has been reported for residents of jails from other US regions (ours: 38.7%, previously reported: 40.9-56.7%).^7–10^

The strength of the Connecticut DOC’s vaccination program is evidenced by the results of our survival analysis among people who become incarcerated following vaccine eligibility. Among this population, we found that following the same amount of time spent eligible for the vaccine, people currently incarcerated were 12.5 times more likely to initiate vaccination than people prior to incarceration. While our findings highlight the ability of within-facility vaccine programs to improve vaccine coverage among this population, the moderate and racially imbalanced vaccine acceptance proportion points to the need for ongoing, evidence-based vaccination program development.

Our results also speak to the need for evidence-based community program development outside of correctional facility settings. This stems from the low level of vaccination among our included residents prior to incarceration. While not examined here, existing data suggests that effective programs should employ accessible and trusted sources of information, such as friends and family, to address vaccine-related concerns and foster trust in medical personnel.^7,8,12^

Our analysis was subject to limitations. First, not all vaccine offers are recorded, and our acceptance numbers are based solely on recorded offers. Second, our vaccination rates are based on recorded doses (from the community or facilities). Third, the DOC racial data is subject to missing data and reporting errors. Finally, our selection of residents and at-risk time for the survival analysis may have introduced bias. While the sensitivity analyses highlight the robustness of our findings, the magnitude of the observed association varied by examined sample.

### Public Health Implications

Despite observing moderate levels of vaccine acceptance among residents of state jails, we found newly incarcerated persons were far more likely to initiate vaccination following incarceration than prior to incarceration. Though our findings highlight the utility of vaccination programs within correctional facilities, they also speak to the need for ongoing, evidence-based vaccination program development within correctional facilities and the community.

## Data Availability

The data used in this study belongs to the Connecticut Department of Correction. Qualified researchers may submit a data share request for de-identified patient level data by contacting the corresponding author with a detailed description of the research question.

## Additional Contributions

We thank the Connecticut Department of Correction and the healthcare workers in each correctional facility for administrative support. None of the individuals received compensation for their contribution.

## Author Contributions

Margaret L. Lind had full access to all the data in the study and takes responsibility for the integrity of the data and the accuracy of the data analysis.

*Concept and design:* Lind, Ko, Kennedy, Richeson

*Acquisition, analysis, or interpretation of data:* Lind, Dorion Nieto, Houde

*Drafting of the manuscript:* Lind, Robertson, Ko, Cummings

*Critical revision of the manuscript for important intellectual content:* All authors.

*Statistical analysis:* Lind

*Administrative, technical, or material support:*

*Supervision:* Ko, Kennedy, Richeson, Cummings

