## Supplement for "Covid-19 Vaccine Acceptance Among People Incarcerated in Connecticut State Jails"

**Supplemental Appendix**

**Vaccination program:** The Connecticut State Department of Correction began their vaccination rollout program on February 2^nd^, 2021. Vaccines were rolled out in alignment with the age-specific guidelines of the state. The age specific rollout dates for the state and DOC were:

| **eTable1: Vaccine Eligibility by Date and Location** | | |
| --- | --- | --- |
|  | **Connecticut** | **Connecticut DOC** |
| ≥ 75 years | 14-Jan-21 | 2-Feb-21 |
| ≥ 65 years | 11-Feb-21 | 11-Feb-21 |
| ≥ 45 years | 19-Mar-21 | 19-Mar-21 |
| ≥ 16 years | 1-Apr-21 | 1-Apr-21 |
| ≥ 12 years^a^ | 13-May-21 | 13-May-21 |
| ^a^DOC facilities house residents aged 15 years or older | | |

| **e.Table2: Recorded First Vaccine Offers and Acceptance Proportion for Residents Incarcerated in a Connecticut State Jail Between February 2nd 2021 and November 8th 2021 by Demographic Characteristics** | | | | |
| --- | --- | --- | --- | --- |
|  | **Total** | **Offered** | **Accepted First Offer** | **Accepted Subsequent Offer** |
|  | **(N=6522)** | **(N=4817)** | **(N=1198)** | **(N=664)** |
| **Demographics** |  |  |  |  |
| Age years, Median (IQR) | 36.0 [18.0, 79.0] | 35.0 [18.0, 79.0] | 39.0 [18.0, 77.0] | 36.0 [18.0, 74.0] |
| Race/Ethnicity, n (%) |  |  |  |  |
| Hispanic/Latinx | 1895 (29.1%) | 1385 (28.8%) | 401 (33.5%) | 210 (31.6%) |
| Non-Hispanic Black | 2596 (39.8%) | 1964 (40.8%) | 391 (32.6%) | 263 (39.6%) |
| Non-Hispanic White | 1979 (30.3%) | 1432 (29.7%) | 395 (33.0%) | 186 (28.0%) |
| Other Race | 52 (0.8%) | 36 (0.7%) | 11 (0.9%) | 5 (0.8%) |
| Incarcerated prior to vaccine program implementation (Feb 2nd 2021) |  |  |  |  |
| First incarceration event prior to Feb 2nd | 2010 (30.8%) | 1899 (39.4%) | 747 (62.4%) | 275 (41.4%) |
| First incarceration event after Feb 2nd prior to age-based eligibility | 772 (11.8%) | 653 (13.6%) | 162 (13.5%) | 111 (16.7%) |
| First incarceration event after Feb 2nd while eligibility^a^ | 3740 (57.3%) | 2265 (47.0%) | 289 (24.1%) | 278 (41.9%) |

***e.Figure1: Population Flowchart for Vaccine Acceptance Examination***

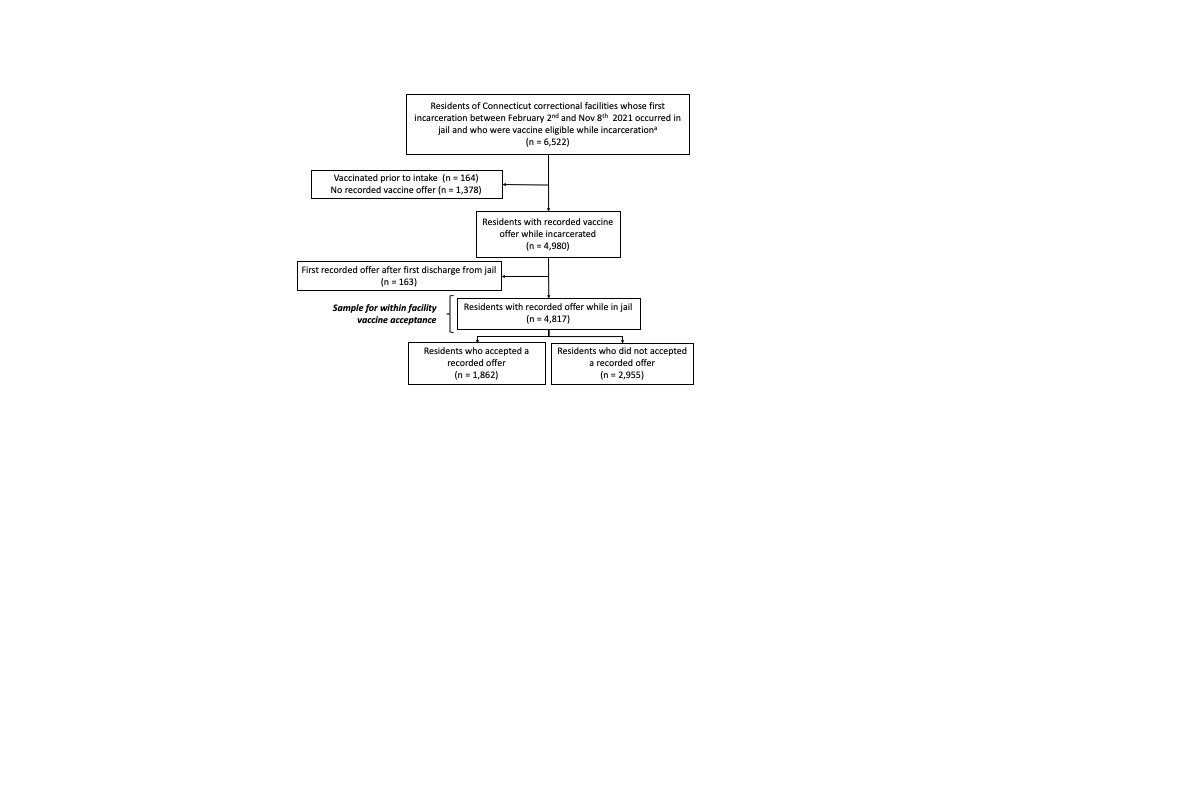

**Time Till Vaccine Initiation Analysis:**

As outlined in the manuscript, our survival analysis was limited to residents who become incarcerated on the day or after they become eligible for the vaccine in the state of Connecticut according to their age.

***e.Figure2: Population Flowchart for Survival Analysis***

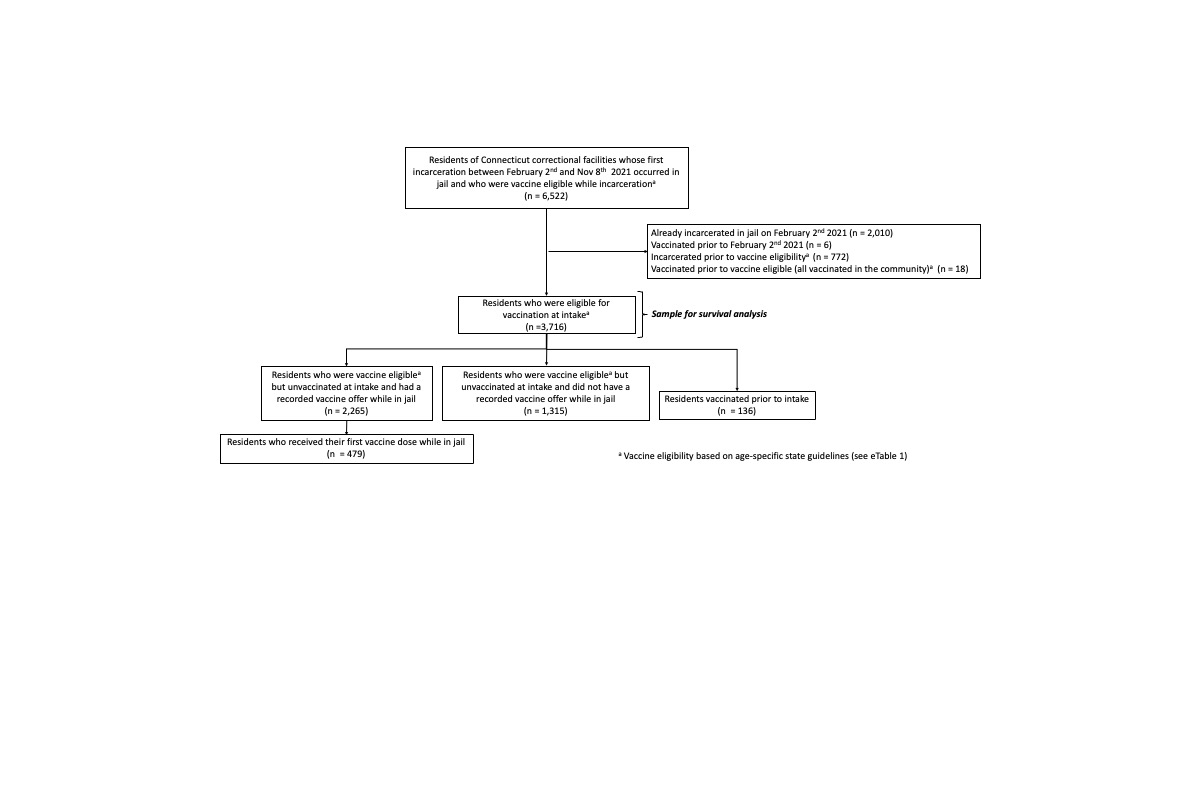

***Proportionality Test:***

We estimated hazard ratios using Cox Proportional Hazards models. To test the proportionality assumption, we tested the slope of the Schoenfeld Residuals with an alpha of 0.05 and graphed the residuals for visual examination. At this alpha, we found no evidence that our proportionality assumption was violated. Below are the Schoenfeld Residual plots and the pvalues for our age and race adjusted as well as our race stratified Cox Proportional Hazard Models.

***e.Figure3: Age and Race Adjusted (All residents)***

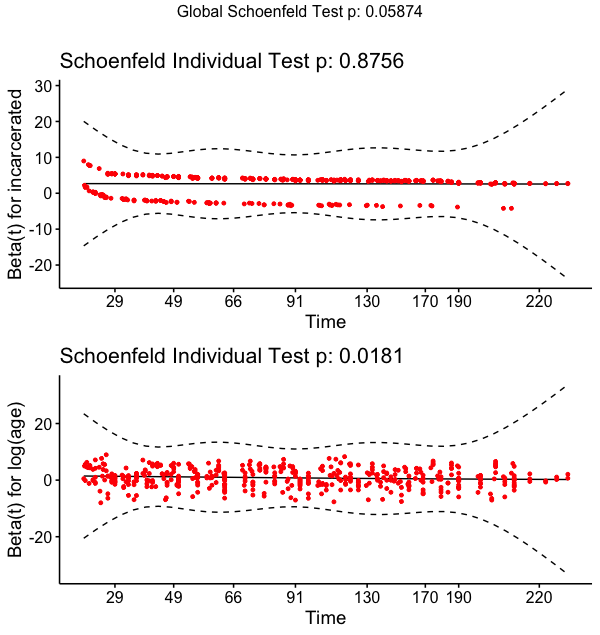

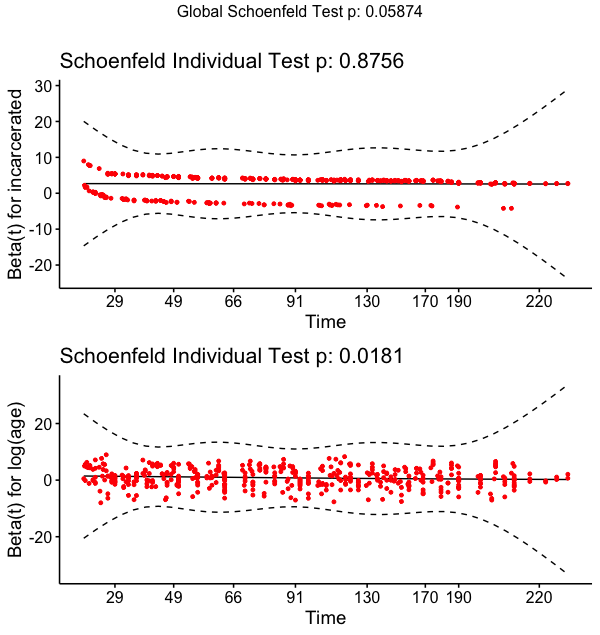

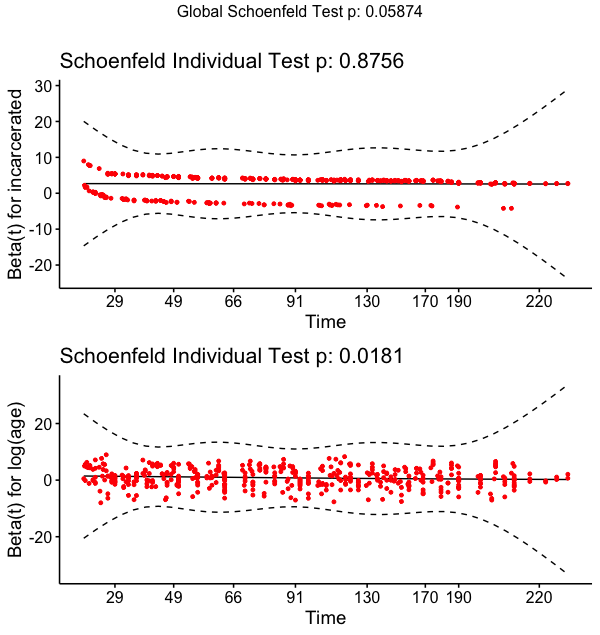

***e.Figure4: Age Adjusted (Non-Hispanic Black)***

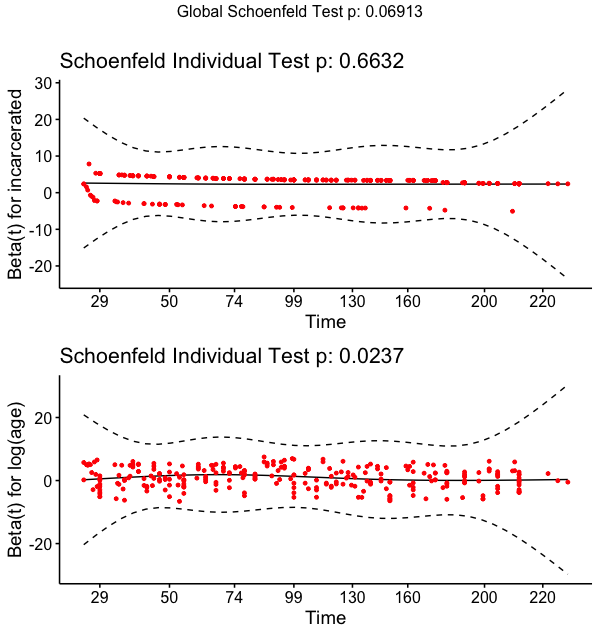

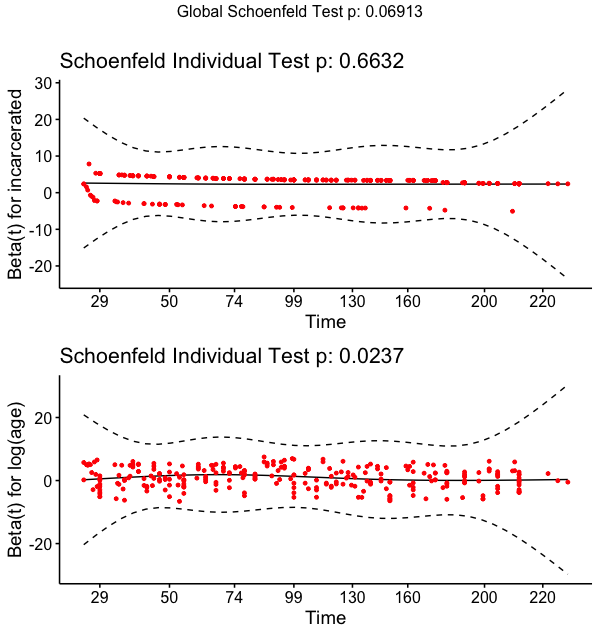

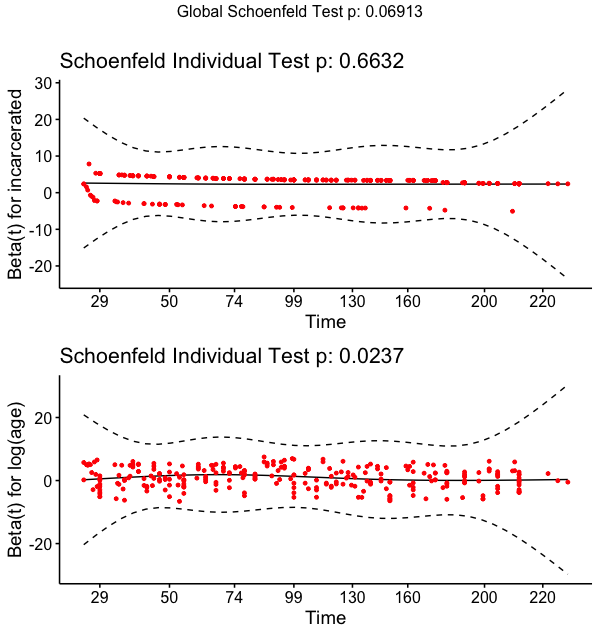

***e.Figure5: Age Adjusted (Non-Hispanic White)***

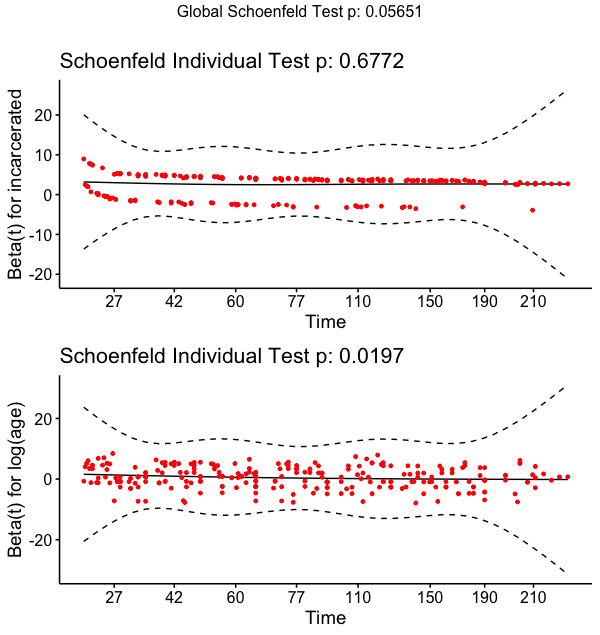

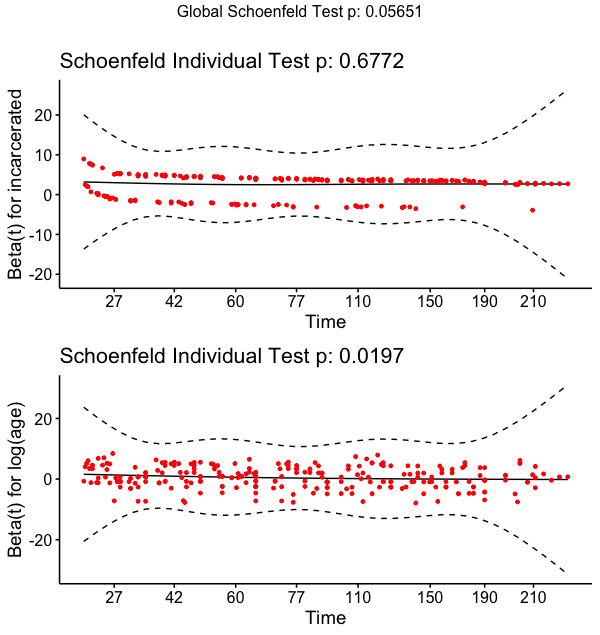

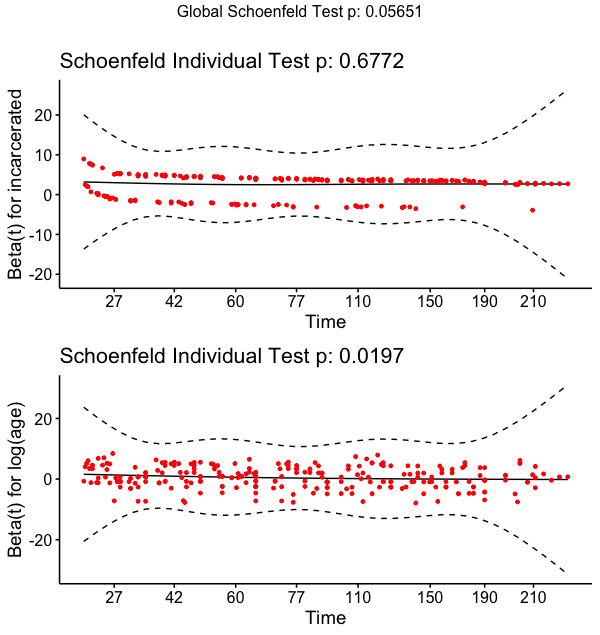

***E.Figure6: Age Adjusted (Hispanic/Latinx)***

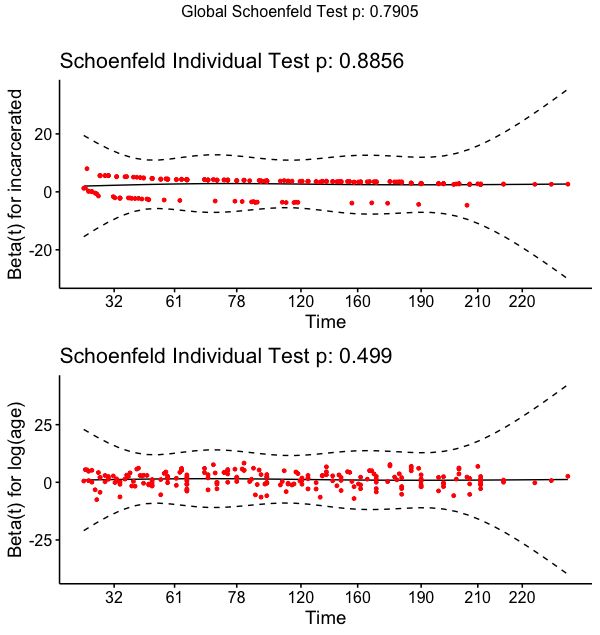

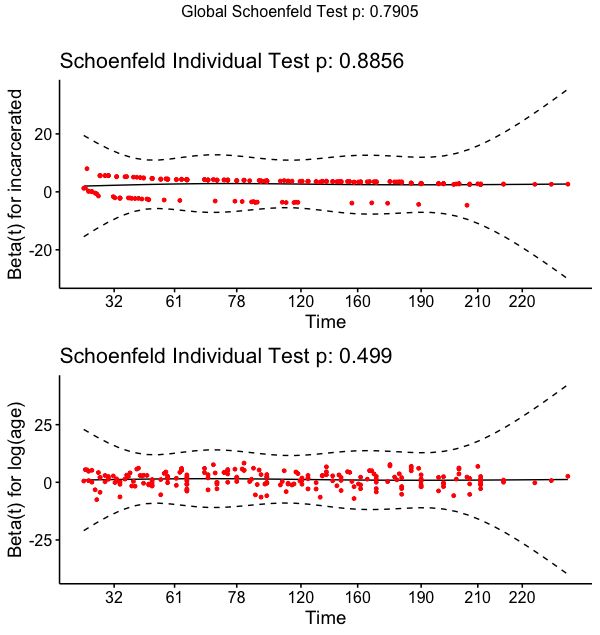

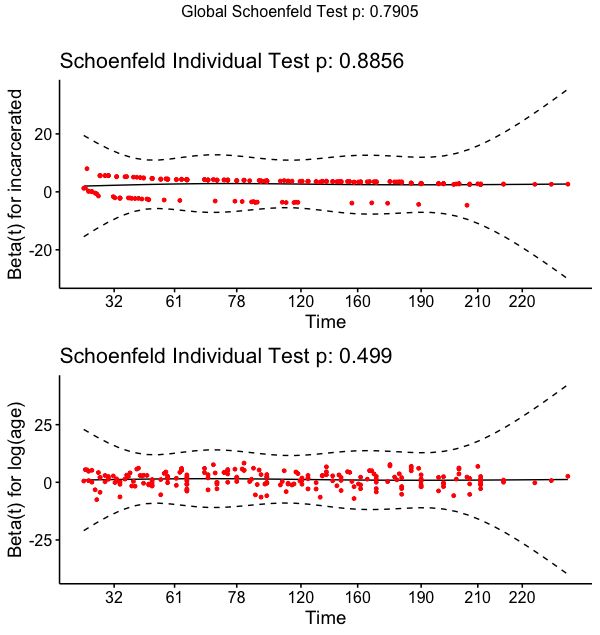

**Demographic Characteristics of Newly Incarcerated Residents of Connecticut State Jails Who Were Vaccine Eligible at Intake by Location of First Vaccine Dose**

***e.Table3: Demographic and Vaccination Characteristics of Residents Incarcerated in Jail on the Day of or Following Statewide, Age-Specific Vaccine Eligibility***

|  | **Total** | **Unvaccinated*** | **Vaccinated in Community** | **Vaccinated in Facility** |
| --- | --- | --- | --- | --- |
|  | **(N=3716)** | **(N=3101)** | **(N=136)** | **(N=479)** |
| **Demographics** |  |  |  |  |
| Age years, Median (IQR) | 35 [28, 45] | 34 [27, 44] | 41 [33, 52] | 38 [32, 48] |
| Race/Ethnicity, n (%) |  |  |  |  |
| Hispanic/Latinx | 1069 (28.8%) | 888 (28.6%) | 40 (29.4%) | 141 (29.4%) |
| Non-Hispanic Black | 1395 (37.5%) | 1173 (37.8%) | 46 (33.8%) | 176 (36.7%) |
| Non-Hispanic White | 1217 (32.8%) | 1012 (32.6%) | 49 (36.0%) | 156 (32.6%) |
| Other Race | 35 (0.9%) | 28 (0.9%) | 1 (0.7%) | 6 (1.3%) |
| **Eligible Time** |  |  |  |  |
| Time in community, median (IQR) | 79 [41, 183] | 90 [44, 185] | 50 [33, 89] | 58 [34, 104] |
| Time in jail, median (IQR) | 14 [3, 31] | 14 [4, 34] | 0 [0, 0] | 10 [5, 31] |
| * Remained unvaccinated as of November 8th, 2021, or upon departure from DOC facility | | | | |

**Sensitivity analysis:**

In our primary analysis comparing time till vaccine initiation in the community to time till vaccination within jails, we included all residents who became incarcerated on the day or after their age made them eligible for vaccination in Connecticut. We chose this population because it allowed us to examine the impact of the vaccine program among people who experienced eligible time for vaccination in both community and jail settings. While we feel this is the best sample from which to conduct this analysis, it is subject to bias. To examine the impact of sample selection bias, we conducted four sensitivity analyses among different variants of the sample. For each analysis, we estimated age and race adjusted hazard ratios.

***Restrict to residents incarcerated on or after May 15^th^*:**

Because vaccine rates within the community increased over time, we were worried that our HRs may overestimate the impact of the vaccine program in later calendar time. In this sensitivity analysis, we limited our sample to those residents with incarceration dates on or after May 15^th^, 2021. As with the primary analysis, we excluded people prior to the start of the analytical period (March 15) but defined start time based on the study period (March 15^th^). This analysis allows us to compare time till vaccination initiation when vaccine update was higher within the community. This restriction resulted in our sample declining to 2,918 residents (102 residents vaccinated in the community and 342 residents vaccinated while incarcerated).

| ***eTable4: Sensitivity Analysis - Adjusted Hazard Ratios Comparing Time Prior to and Following Incarceration in Connecticut State Jail (limited to residents who were incarcerated on or after May 1st 2021)*** | | |
| --- | --- | --- |
|  | **HR** | **95% CI** |
| All Residents | 22.2 | (16.7, 29.5) |
| Race/Ethnicity |  |  |
| Non-Hispanic Black | 18.6 | (11.6, 30.1) |
| Non-Hispanic White | 20.0 | (12.5, 32.2) |
| Hispanic/Latinx | 28.0 | (16.3, 47.1) |
| * Estimated using Cox Proportional Hazard Models adjusted for age | | |

***Restrict to residents offered vaccines*:**

In the primary analysis, we included all residents who were eligible for vaccination upon intake in our sample. However, not all unvaccinated residents were offered a vaccine during their incarceration or prior to the end of the study period and our HRs may underestimate the impact of the vaccine program if universal vaccination was provided. While we chose to look at the program in its existing state (as of November 2021), here, we limit the sample to people vaccinated in the community or offered a vaccine following incarceration. This resulted in a sample of 2,401 (136 residents vaccinated in the community and 479 vaccinated while incarcerated).

| ***eTable5: Sensitivity Analysis - Adjusted Hazard Ratios Comparing Time Prior to and Following Incarceration in Connecticut State Jail (limited to community vaccine or facility offered residents)*** | | |
| --- | --- | --- |
|  | **HR** | **95% CI** |
| All Residents | 7.0 | (5.6, 8.6) |
| Race/Ethnicity |  |  |
| Non-Hispanic Black | 5.5 | (3.8, 7.8) |
| Non-Hispanic White | 8.9 | (6.2, 12.6) |
| Hispanic/Latinx | 6.9 | (4.6, 10.3) |
| * Estimated using Cox Proportional Hazard Models adjusted for age | | |

***Include all community eligibility time and vaccination events*:**

To increase the comparability between the unexposed and exposed follow-up time, we limited our primary analysis to incarceration and vaccination events occurring on or after the Connecticut DOC began their vaccination program (February 2^nd^ 2021). This removed almost a month of eligible vaccination time from residents aged ≥75 years in the community. Here, we expanded our eligibility time in the community to start on January 14^th^ for residents ≥75 years of age. However, no ≥75-year-olds incarcerated on or after February 2^nd^, 2021 were vaccinated during this expanded period. For this reason, this expanded exposure period only led to expanded “at risk” time in the community for the four residents ≥75 years old. This was limited as only six residents met this criterion.

| ***eTable6: Sensitivity Analysis - Adjusted Hazard Ratios Comparing Time Prior to and Following Incarceration in Connecticut State Jail (all community vaccination and eligibility time including pre-February 2nd, 2021)*** | | |
| --- | --- | --- |
|  | **HR** | **95% CI** |
| All Residents | 12.9 | (10.5, 15.8) |
| Race/Ethnicity |  |  |
| Non-Hispanic Black | 10.9 | (7.7, 15.4) |
| Non-Hispanic White | 14.2 | (10.1, 20.0) |
| Hispanic/Latinx | 13.9 | (9.4, 20.3) |
| * Estimated using Cox Proportional Hazard Models adjusted for age | | |

***Restrict to people who contributed at least a week of eligible time in the community:***

In our primary analysis, we included residents who became incarcerated on the day they become eligible for vaccination. Because these residents have a very low probability of being vaccinated in the community prior to incarceration, we wanted to ensure that their inclusion was not driving the analysis. To do so, we restricted our sample to people who were eligible for vaccination within the community for at least one week prior to incarceration. This restriction resulted in our sample declining to 3596 residents (136 residents vaccinated in the community and 460 residents vaccinated while incarcerated).

| ***eTable7: Sensitivity Analysis - Adjusted Hazard Ratios Comparing Time Prior to and Following Incarceration in Connecticut State Jail (limited to residents who were eligible for the vaccine for at least 7 days prior to incarceration)*** | | |
| --- | --- | --- |
|  | **HR** | **95% CI** |
| All Residents | 12.8 | (10.4, 15.8) |
| Race/Ethnicity |  |  |
| Non-Hispanic Black | 11.3 | (8.0, 16.1) |
| Non-Hispanic White | 13.3 | (9.4, 18.7) |
| Hispanic/Latinx | 14.2 | (9.6, 21.0) |
| * Estimated using Cox Proportional Hazard Models adjusted for age | | |
